# Radiomics with super-voxel segmentation improves HPV prediction accuracy significantly in oropharyngeal cancer

**DOI:** 10.1101/2024.12.23.24319468

**Authors:** Oya Altinok, Matthew B. Schabath, Albert Guvenis

**Affiliations:** Institute of Biomedical Engineering, Bogazici University, Istanbul, Turkiye; Department of Cancer Epidemiology, H. Lee Moffitt Cancer Center & Research Institute, Tampa, FL, USA; Biomedical Engineering, Namik Kemal University, Tekirdag, Turkiye

**Keywords:** Sub-region analysis, Radiomics, Oropharyngeal cancer

## Abstract

Human papillomavirus (HPV) status has been shown to be prognostic among patients with oropharyngeal cancer (OPC); Specifically, patients with HPV-positive tumors often have a superior prognosis and response to treatment compared to HPV-negative tumors which may be attributed greater tumor heterogeneity. This study assessed if analyzing tumor subregions (i.e., habitats) using super-voxel segmentation can effectively capture intratumor heterogeneity and improve predicting HPV positivity compared to a conventional whole-tumor approach. Using publicly available data from The Cancer Imaging Archive (TCIA) of 192 patients (85% HPV positive) with OPC, we utilized radiomics to predict HPV status comparing a super-voxel segmentation approach and a whole tumor approach. For the subregion approach, the number of supervoxels (subregions) generated per patient varied based on tumor size (mean = 30 supervoxels/patient [SD = 10]). 18 radiomic features were extracted from each supervoxel based on gray-level frequency distribution and aggregated using variance to summarize heterogeneity. For the whole tumor approach, the same radiomic features were generated across the entire tumor without sub-segmentation. As such, 18 radiomic features were utilized to predict HPV status in both models. The dataset was divided into a training set (70%) and an independent test set (30%). An optimizable ensemble model based on a decision tree with GentleBoost was applied to both the subregion and the whole tumor models to predict HPV status from radiomics features. The proposed super-voxel-based approach yielded an AUC of 0.94 in the training set and 0.91 in the test set which outperformed whole tumor analysis (AUC of 0.77 and 0.75, respectively). These findings demonstrate the value of incorporating heterogeneity measures and super-voxel segmentation in oropharyngeal cancer radiomics, which enable a significantly more accurate prediction of the HPV Status.

## 1. INTRODUCTION

Tumors are comprised of heterogenous and diverse cell populations at the genetic, epigenetic, proteomic, and phenotypic levels [1–3]. Tumor heterogeneity among oropharyngeal cancers (OPC) is well characterized [4] and this complexity is further influenced by various exogenous risk factors such as smoking, alcohol, oral hygiene, diet, genetics, and HPV status. HPV-positive tumors often have a better prognosis and response to treatment compared to HPV-negative tumors [5,6], which may exhibit a higher level of heterogeneity [7–9]. Increased levels of tumor heterogeneity have been reported to be associated with poor clinical outcomes in OPC [4]. As standard-of-care in the clinical setting, a single-point biopsy is taken for cancer diagnosis, and this may not be sufficient to explain the genetic profile of the entire tumor [10]. There approaches to capture tumor heterogeneity, such as evaluating multiple samples from individual tumors [11] or performing single-cell analyses [12]. However, these methods are invasive, require tissues, and cannot be calculated rapidly. Conversely, image- and radiomics-based methods are non-invasive, can rapidly identify subregions of tumors from standard-of-care medical images, and can capture data from the entire region of interest rather than a small portion of the tumor that is biopsied.

As such, there is an emerging and critical need to develop non-invasive and rapidly measured biomarkers in clinical care. However, there is a significant gap in current radiomics approaches as they largely ignore heterogeneity (reviewed in [13]). For instance, features extracted from an image may represent the average of these different cell types, potentially obscuring important information about specific subpopulations within the tumor. Therefore, accounting for and analyzing this heterogeneity in radiomics studies may be important for developing accurate and personalized cancer diagnosis and treatment strategies in OPC [4,14].

We propose enhancing HPV prediction accuracy in radiomic studies by incorporating "heterogeneity measurements". Measures of heterogeneity can be an additional factor in predicting HPV status in OPC. Our previous HPV prediction model [15] showed acceptable accuracy. We believe that the model’s accuracy will be improved by considering subregion radiomics as demonstrated in studies [16–18]. This information is likely to change the therapeutic plan. There are studies in the literature (i.e., [19]).

The focus of this study was on extracting first-order features due to their simplicity, interpretability, and ability to provide a direct measure of overall tumor characteristics [20].

Features like skewness, kurtosis, and variance provide valuable insights into the distribution of voxel intensities, which is particularly useful when comparing HPV-positive and HPV-negative tumors [21].

## 2. MATERIALS AND METHODS

### 2.1. Data Sets

The dataset used in this analysis from The Cancer Imaging Archive (TCIA) consists of 192 patients (**Table 1**) diagnosed with OPC and includes pre-treatment, contrast-enhanced CT images, segmentation labels retrieved [22–24], and HPV status (p16 protein-positive or negative). The gross primary tumor volume (GTVp), segmented by experts [23], was considered in the radiomic analysis. Initially, there were 412 patients, but after excluding 145 patients with missing HPV status and 75 patients with segmentation errors, 192 patients remained for analysis.

**Table 1.**
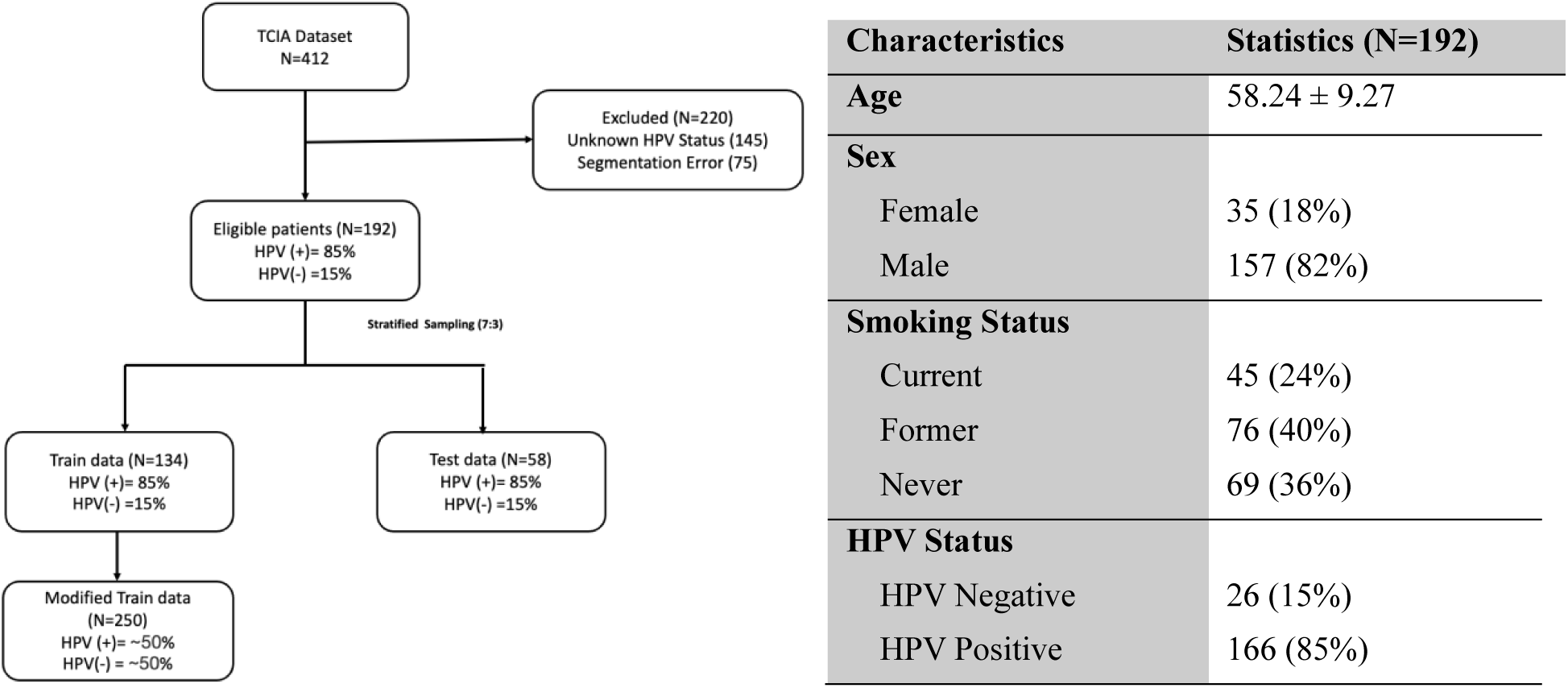
Patient characteristics of all included patients. Continuous variables are represented as means ± standard deviation. The diagram illustrates patient selection. Dataset were retrieved from The Cancer Imaging Archive (TCIA) [22–24].

### 2.2. Imaging Preprocessing

In the imaging preprocessing step, all CT scans were resampled to a uniform voxel size of 1 mm × 1 mm × 1 mm using cubic interpolation to address inconsistencies in the original voxel sizes. After resampling, z-score normalization was applied to standardize the data. Next, entropy maps were generated and applied to the CTs. The original CT images and the entropy-filtered CTs were then fused for further analysis. Resampling, interpolation, and preprocessing were performed using MATLAB 2023a.

### 2.3. Measuring Tumor Heterogeneity

The objective of this analysis to determine whether measures of tumor heterogeneity can be incorporated as an additional factor in predicting HPV status in OPC. **Figure 1** presents the workflow of our study. We compared two approaches for predicting outcomes from radiomics features extracted from CT scans of OPC patients: super-voxel segmentation approach versus a whole tumor approach

**Figure 1.**
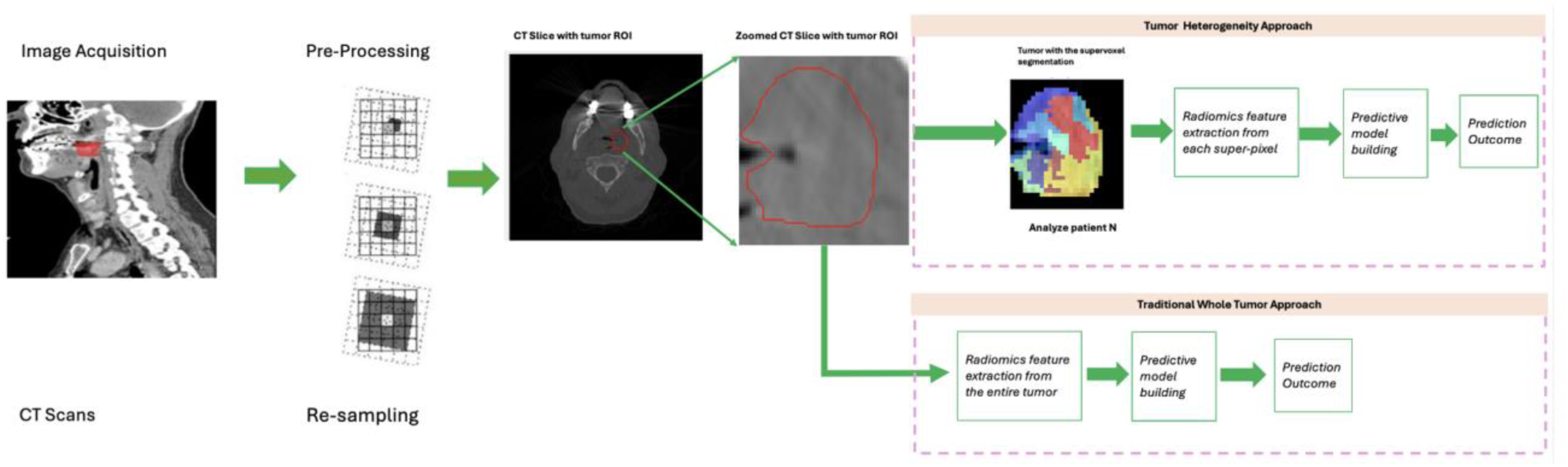
Illustration of Tumor Heterogeneity Approach and Traditional Whole Tumor Approach Workflow. The figure shows a CT slice with the tumor region highlighted by a red contour. For illustration of this process, 2D images are used.

#### 2.3.1. Super-Voxel Segmentation

Tumors were over-segmented into super-voxels at the patient level using the SLIC (Simple Linear Iterative Clustering) algorithm, with Euclidean distance as the similarity metric. For the super- voxel approach, we adapted the method from reference [25]. The fused image, containing both intensity information from CT images and texture information from entropy maps, was used as input for the segmentation process. Combining entropy values with intensity-based CT features creates a richer dataset [26].

The number of super-voxels generated was dynamically adjusted based on the size of the tumor mask. Each super-voxels were assigned a label based on similarity in both intensity and spatial proximity, ensuring that regions with similar characteristics were grouped together. Supervoxel analysis was performed in MATLAB 2023a using the *superpixels3* function.

#### 2.3.2. Subregion Feature Extraction and Feature Aggregation

To capture patient-level intrinsic heterogeneity of subregions, we applied feature aggregation (**Figure 2**) using variance analysis to the radiomics features extracted from subregions (supervoxels) to summarize heterogeneity across the tumor regions. **Figure 2** illustrates a three-stage process used for that. Each stage’s detail is given as follows.

**Figure 2.**
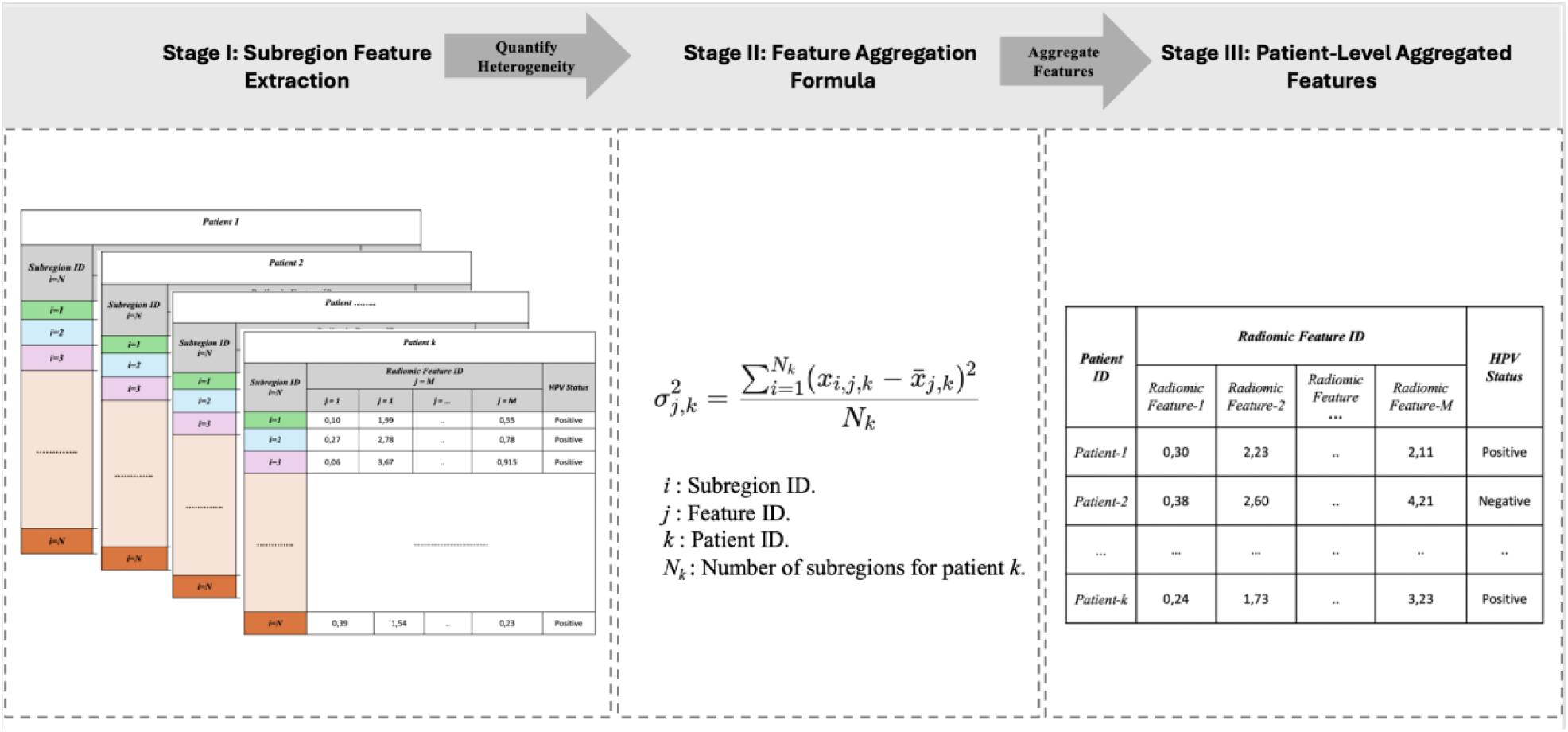
Workflow for subregion feature extraction, feature aggregation, and patient level feature analysis. The figure illustrates a three-stage process for features used for tumor heterogeneity.

### Stage I: Subregion Feature Extraction

Radiomic features are extracted from tumor subregions (denoted as *i=1,2,…, N_k_*) for each patient. Each subregion is uniquely identified (e.g., *i*). Multiple radiomic features *(j=1,2,…, M*) are calculated for each subregion. The table in this panel demonstrates the layout of extracted features for subregions from individual patients.

Extracted radiomic feature names are given in **Table 2**. These features were computed separately for both the original CT images and the entropy-filtered CT images. First-order features (n = 18) were derived from the gray-level frequency distribution using histogram analysis for each super- voxel. Feature extraction was performed using MATLAB 2023a.

**Table 2.** Aggregated Radiomic Features Extracted from CT and Entropy Filtered CT Images of Supervoxels.

| **Table.2** Aggregated Radiomic Features Extracted from CT and Entropy Filtered CT Images of Supravoxels |  |  |  |
| --- | --- | --- | --- |
| **Feature Type** | **Feature Aggregation** | **CT Feature Name** | **Entropy Filtered CT Feature Name** |
| Skewness | Variance | Var_Skewness_1 | Var_Skewness_2 |
| Kurtosis | Variance | Var_Kurtosis_1 | Var_Kurtosis_2 |
| Mean | Variance | Var_Mean_1 | Var_Mean_2 |
| Median | Variance | Var_Median_1 | Var_Median_2 |
| First Quartile (25th) | Variance | Var_Quantile25_1 | Var_Quantile25_2 |
| Third Quartile (75th) | Variance | Var_Quantile75_1 | Var_Quantile75_2 |
| Interquartile Range | Variance | Var_IQR_1 | Var_IQR_2 |
| Variance | Variance | Var_Variance_1 | Var_Variance_2 |
| Energy | Variance | Var_Energy_1 | Var_Energy_2 |

### Stage II: Statistical Aggregation Formula

To quantify tumor heterogeneity, the variance 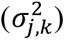 of each radiomic feature (*j*) is computed across all subregions (*i*) within a given patient (*k*). The formula for variance is provided:

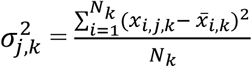

where 𝑥_𝑖,𝑗,𝑘_ is the value of *j* -th feature for the *i* -th subregion of the *k*-th patient, 𝑥̅_𝑖,𝑘_ is the mean value of the *j* -th feature across all subregions, and 𝑁_𝑘_ is the total number of subregions for the *k*-th patient. This step allows for patient-level feature aggregation.

### Stage III: Patient-Level Aggregated Features

The aggregate variance values 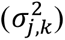 for each radiomic feature (*j*) are organized into a patient-level table. Each row corresponds to a patient (*k*), with columns representing the aggregated features (*j* = 1, 2, . . ., *M*) and the associated HPV status (Positive or Negative). These patient-level features serve as input for downstream machine learning analyses to predict HPV status and quantify tumor heterogeneity.

This approach was crucial for assessing heterogeneity within the tumor, as the variance of super-voxel features reflects how much the tumor structure differs across different regions. By summarizing the radiomic features using variance, we were able to capture the internal diversity of the tumor in a more granular way compared to traditional whole-tumor feature extraction, which may overlook these subtle differences. Additionally, this aggregation reduced the dimensionality of the dataset derived from the super-voxel radiomic feature set, simplifying the analysis while preserving the essential information about tumor heterogeneity.

#### 2.3.3. Data Preprocessing

After feature extraction, median imputation was applied to handle missing values. Next, z-score normalization was performed to standardize the data. This preprocessing was done in MATLAB 2023a. The same process was followed for both the features extracted from super-voxels and those extracted from the whole tumor.

#### 2.3.4. Data Splitting and Balancing

The dataset was divided into training and test sets using stratified sampling to ensure an even distribution of HPV status across both sets. Specifically, 70% of the data was used for training, while the remaining 30% of the data was kept as independent test data.

To address class imbalance, Synthetic Minority Oversampling Technique (SMOTE) was applied to the training data. Synthetic samples were generated by interpolating between k-nearest neighbors of the minority class to achieve a balanced class distribution.

### 2.4. Traditional Whole Tumor Approach

To ensure comparability with the super-voxel approach, entropy-filtered CT images were included alongside the original CT for the whole tumor analysis. Features were calculated across the entire tumor region using both the original and entropy-filtered CTs. Extracted features, shown in Table 3, include skewness, kurtosis, mean, median, first quartile, second quartile, interquartile range, variance, and energy. These features were computed separately for the original and entropy-filtered CT images using MATLAB 2023a.

**Table 3:** Radiomic Features Extracted from Whole Tumor Analysis.

| Table 3: Radiomic Features Extracted from Whole Tumor Analysis |  |  |
| --- | --- | --- |
| Feature Type | CT Feature Name | Entropy Filtered CT Feature Name |
| Skewness | Skewness 1 | Skewness 2 |
| Kurtosis | Kurtosis 1 | Kurtosis 2 |
| Mean | Mean 1 | Mean 2 |
| Median | Median 1 | Median 2 |
| First Quartile (25th) | Quantile25_1 | Quantile25_2 |
| Third Quartile (75th) | Quantile75_1 | Quantile75_2 |
| Interquartile Range | IQR_1 | IQR_2 |
| Variance | Variance_1 | Variance_2 |
| Energy | Energy_1 | Energy_2 |

#### 2.4.1 Data Preprocessing for Whole Tumor Approach

After feature extraction, median imputation was applied to handle missing values. Next, z-score normalization was performed to standardize the data. This preprocessing was done in MATLAB 2023a. Data preprocessing was applied to the features extracted from the whole tumor.

#### 2.4.2. Data Splitting and Data Balancing

The dataset was divided into training and test sets using stratified sampling to ensure an even distribution of HPV status across both sets. Specifically, 70% of the data was used for training, while the remaining 30% of the data was kept as independent test data.

To address class imbalance, Synthetic Minority Oversampling Technique (SMOTE) was applied to the training data. Synthetic samples were generated by interpolating between k-nearest neighbors of the minority class to achieve a balanced class distribution.

### 2.5. Model Building and Performance Evaluation

To predict HPV status, an optimizable ensemble model was built using MATLAB 2024b Classification Learner. The model used a decision tree as the base learner, and hyperparameters were optimized using GentleBoost. This model was chosen due to its effectiveness in handling moderate feature sets (in this case, 18 features) and its ability to boost performance by combining multiple weak learners to reduce bias and variance, making it suitable for our dataset [27]. To compare the whole tumor-based radiomics model, the same model construction was applied to build the whole tumor HPV prediction model. The aggregated features (Table 2) for the tumor heterogeneity approach were used, while for the whole tumor approach, the features extracted from the entire tumor were used (Table 3).

### 2.6. Hyperparameter Tuning

Selecting appropriate hyperparameters in machine learning is crucial for improving classifier accuracy and overall model performance [28]. Fine-tuning these hyperparameters can significantly impact the effectiveness of a model by optimizing its behavior for the given data. Our study focused on optimizing key hyperparameters, including the learning rate, number of learners, and maximum number of splits, for the selected ensemble classifier (GentleBoost).

We employed Bayesian optimization, a systematic approach that efficiently explores the hyperparameter space to identify an optimal combination. Overfitting is a common challenge in machine learning, where a model performs well on training data but poorly on unseen data. To address this issue and ensure robustness, we applied a 10-fold cross-validation method during optimization. This approach divides the training data into ten folds, iteratively training and validating the model on different folds to minimize bias and variance in performance estimates.

Combining Bayesian optimization with 10-fold cross-validation, we systematically fine-tuned the hyperparameters while mitigating overfitting. The final optimized hyperparameters for the GentleBoost classifier were as follows: the number of learners was 163, the learning rate was 0.4437, and the maximum number of splits was 11. These hyperparameters resulted in a well-performing and robust ensemble classifier.

### 2.7. Model Performance

The predictive performance of the models was assessed using receiver operating characteristic (ROC) curve analysis and the area under the curve (AUC). These metrics were evaluated through a 10-fold cross-validation scheme to ensure robustness and accuracy in the predictions. We employed 10 fold cross-validation (CV=10) on training data (70 %). The remaining 30 % of the data was excluded and kept for testing purposes.

## 3. RESULTS

Overall, the super-voxel method outperforms the whole tumor approach to predict HPV status from CT images. For the super-voxel model, the area under the curve (AUC) was 0.94 for the training set and 0.91 for the test set **(Figure 3),** while for the whole tumor model, the AUC was 0.77 for the training set and 0.75 for the test set **(Figure 3)**.

**Figure 3.**
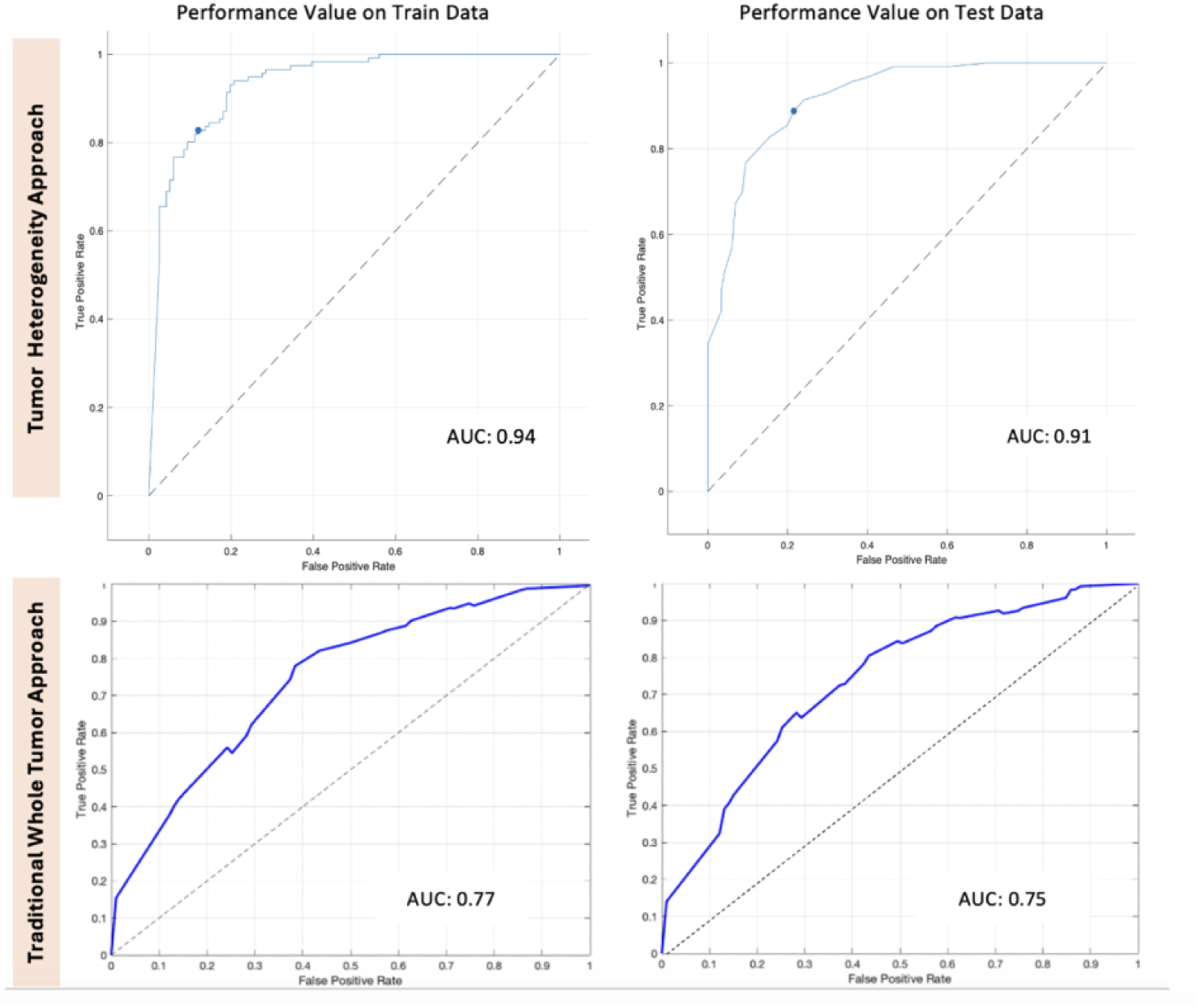
ROC curves and corresponding AUC values are shown for both training and test data.

The number of subregions (or supervoxels) for each tumor varies depending on the tumor’s size and shape. In our study, we observed approximately 30 supervoxels per patient (SD = 10). For example, HPV-positive patients had an average of 30 ± 9 supervoxels, while HPV-negative patients had an average of 30 ± 13 supervoxels.

**Figure 4** further supports this finding by illustrating the differences in tumor heterogeneity between HPV-positive and HPV-negative patients. The distinct regions of variability captured in the super-voxel analysis emphasize the spatial heterogeneity present within the tumor. The bar chart in the bottom row of **Figure 4** compares the mean ’Var_Variance_1’ values between HPV-positive and HPV-negative tumors. The significantly higher variance observed in HPV-negative tumors (*p-value* = 0.025155) indicates greater intratumor heterogeneity, which aligns with their more aggressive clinical behavior. This heterogeneity is a critical factor that may contribute to the improved predictive performance observed with the super-voxel model, as it allows for more nuanced characterization of tumor subregions.

**Figure 4.**
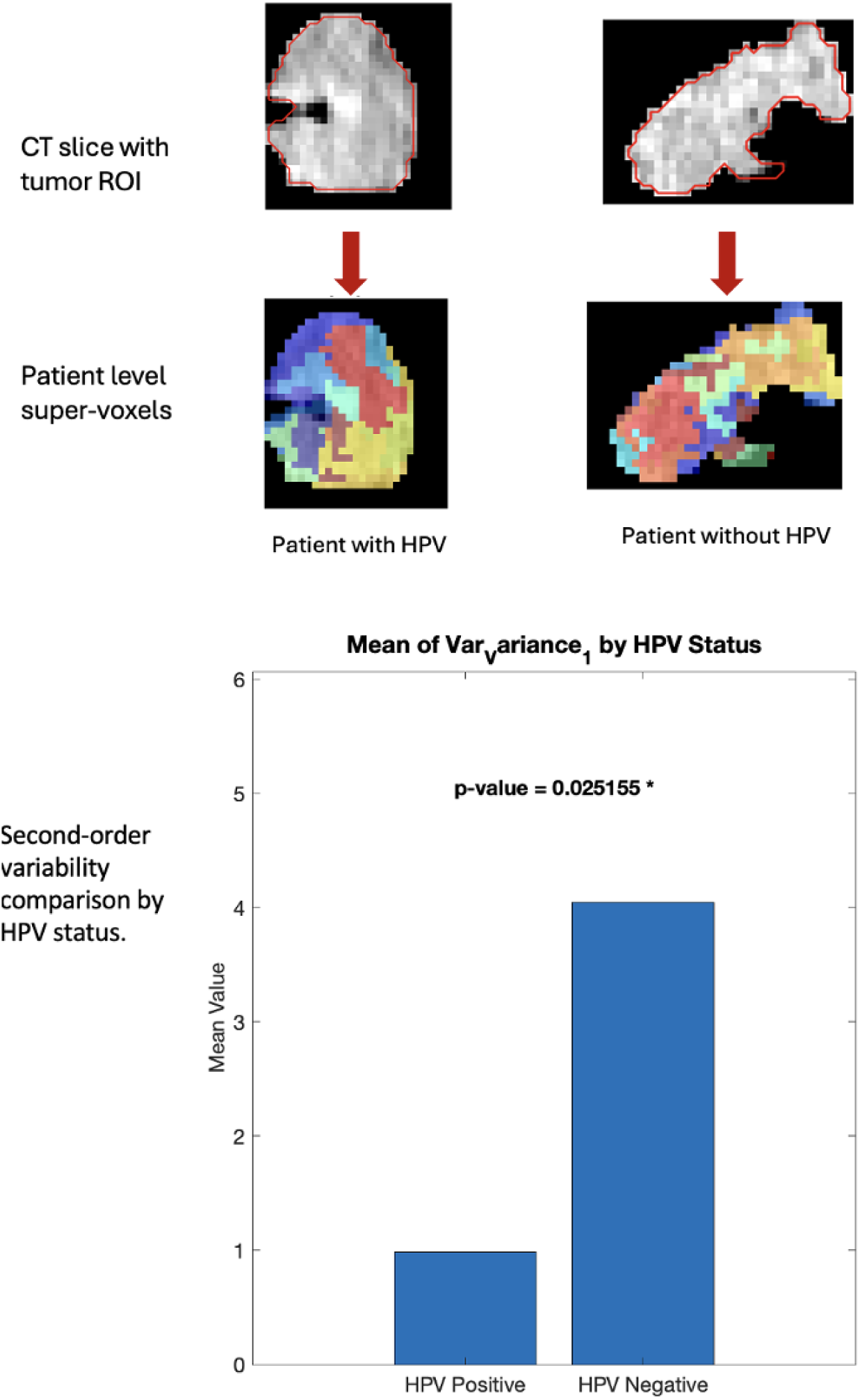
<u>Top Row:</u> CT slices with the outlined tumor region of interest (ROI) for two patients, one with HPV (left) and one without HPV (right). <u>Middle Row:</u> The corresponding super-voxel segmentation of the tumor, with different colors representing distinct regions of variability. The left image shows the super-voxels for a patient with HPV, while the right image shows a patient without HPV. <u>Bottom Plot:</u> Bar chart comparing the mean value of the feature ’Var_Variance_1’ between HPV-positive and HPV-negative tumors. ’Var_Variance_1’ measures the variability of variance within supervoxels across the tumor, reflecting intratumor heterogeneity. The significantly higher variance observed in HPV-negative tumors (p-value = 0.025155, denoted by *) suggests that these tumors exhibit greater heterogeneity compared to HPV-positive tumors.

We quantified the second-order variability of tumor intensity variance across subregions using the feature Var_Variance_1. Each tumor was segmented into supervoxels, and the intensity variance 𝑉𝑖 was calculated for each supervoxel (subregion). Subsequently, the variability among these variance values was computed within the tumor. The calculation involved the following steps:

1. Variance (𝑉𝑖) of the *i* -th Subregion:

The variance of intensity values within each supervoxel (𝑉𝑖) is calculated as:

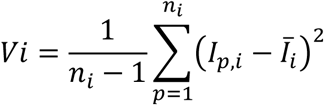

where:

𝐼_𝑝,𝑖_ represents the intensity of the *p* -th voxell in the *i* -th supervoxel.

𝐼̅_𝑖_ is the mean intensity of all voxels the *i* -th supervoxel, calculated as:

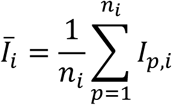

𝑛_𝑖_ is the total number of voxels in the *i* -th supervoxel,

1. 2. Mean Variance (𝑉̅) Across Supervoxels:

The mean variance across all 𝑁 supervoxels in the tumor is computed as:

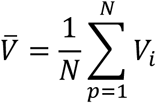

where 𝑁 is the total number of supervoxels (subregions) in the tumor.

1. 3. Second-Order Variability:

Finally, 𝑉𝑎𝑟_𝑉𝑎𝑟𝑖𝑎𝑛𝑐𝑒_1 is calculated to capture the second-order variability of intensity variance across supervoxels:

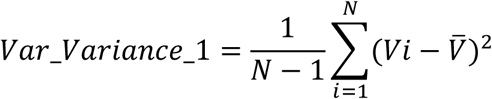

where *Vi* is the variance of the *i* -th supervoxel and 𝑉̅ is the mean variance across all supervoxels. This feature captures the heterogeneity in intensity variance among different tumor regions, reflecting intratumoral heterogeneity. Higher values of 𝑉𝑎𝑟_𝑉𝑎𝑟𝑖𝑎𝑛𝑐𝑒_1 is indicate greater variability in the intensity variance across supervoxels, which may be associated with distinct biological characteristics of the tumor.

## 4. DISCUSSION

In this study we assessed whether incorporating heterogeneity measures into prediction models would improve their performance to predict HPV status from CT images. Using super-voxel segmentation and variance-based aggregation, we were able to capture subtle changes that helped distinguish between HPV-positive and HPV-negative cases.

Although subregion analysis is well-studied for brain and thorax tumors, it is much less explored in head and neck cancer, with only 8% of studies focused on this area (reviewed in reference [29]). Head and neck cancers (HNC) refer to malignancies in regions such as the paranasal sinuses, nasal cavity, oral cavity, tongue, salivary glands, larynx, nasopharynx, oropharynx, and hypopharynx. Given the limited studies focused exclusively on OPC, **Table-4** summarizes tumor subregion analyses HNC, including both OPC and other HNC sites.

**Table 4.** Summary of Tumor subregion analysis for head and neck cancer (HNC). Abbreviations: OS, overall survival; PFS, progression free survival; HPV, Human papillomavirus.

| Reference/Year | Clustering approach | Models | Task | Modality | # of patients in training/testing datasets | Validation Method on test data | Head and Neck Cancer Site |
| --- | --- | --- | --- | --- | --- | --- | --- |
| Wu et al.,2020 [26] | Population level clustering | Cox regression model | Assess early response and predict PFS | <sup>18</sup>F-FDG PET/CT | 81/81 | C-index = 0.66 | Oropharyngeal cancer |
| Xu et al., 2020 [30] | Population level clustering | LASSO | Predict PFS | <sup>18</sup>F-FDG PET/CT | 85/43 | C-index = 0.69 | Nasopharyngeal cancer |
| Pan et al., 2023 [17] | Individual level clustering | Multiple instance learning | Predict local regional recurrence | <sup>18</sup>F-FDG PET/CT | 130/98 | C-index = 0.721 | HNC (includes all tumor sites) |
| Our study | Individual level clustering | Optimizable ensemble model, based on a decision tree with GentleBoost | Predicting HPV Status | CT | 134/58 | AUC = 0.91 | Oropharyngeal cancer |

Most research to date has used population-level subregions, clustering voxel features across different patients, in HNC [26,30] as well as in other cancers [31–33]. These population-level subregions assume uniform tumor phenotypes across patients. However, only one prior study applied individual-level subregion analysis in HNC covering all HNC sites rather than OPC alone [17].

To the best of our knowledge, no individual-level clustering studies have been conducted specifically on OPC. Therefore, our study introduces an individual-level clustering approach with a key innovation: aggregating features across super-voxels by calculating their variance.

One key innovation in our approach was feature aggregation using variance quantification across supervoxels. This method captures tumor heterogeneity while reducing dataset dimensionality. Aggregating features at the supervoxel level simplifies the model but preserves essential information about tumor variations. This allows the model to identify differences across subregions within the tumor. Unlike whole-tumor analysis, which averages features and masks intra-tumoral differences, our approach provides a deeper understanding of tumor complexity.

In the context of predicting HPV status, **Table-5** summarizes relevant studies with diverse methods and features. Bagher-Ebadian et al. [34] utilized a General Linear Model with 12 features, achieving a test AUC of 0.869. Bogowicz et al. [35] applied multivariable logistic regression with 4 features, resulting in a test AUC of 0.78. In our earlier study [15], whole-tumor analysis with Bayesian Networks achieved a test AUC of 0.72. Kubra et. al. [36] used a Random Forest model an achieved a test AUC of 0.77 with 25 features.

**Table 5:** Machine learning models for HPV status prediction in CT of head and neck cancer patients. N represents the total number of features used in the model.

| Reference | Number of patients | Classifier | Performance (AUC) |  | Features |
| --- | --- | --- | --- | --- | --- |
|  |  |  | Train | Test |  |
| Bagher-Ebadian et al.[34] | 187 oropharyngeal cancer patients | General Linear Model | 0.849 | 0.869 | (N=12 features) GLCM – Contrast, Energy,Entropy<br>Shape Features -MeanBreadth & SphericalDisproportion<br>DOST- Energy1,2,3,4,6,7,8,9 |
| Bogowicz et al. [35] | 149 Head and Neck cancer patients | Multivariable logistic regression | 0.85 | 0.78 | (N=4 features) LLL standard deviation, LLL small zone high gray-level emphasis, HHL difference entropy, Coefficient of variation |
| Altinok et al. [15] | 246 oropharyngeal cancer patients | Bayesian Networks | 0.78 | 0.72 | (N=2 features) sphericity and max2DDiameterRow |
| Kubra et. al. [36] | 238 oropharyngeal cancer patients | Random Forest | N/A | 0.77 | (N=25 features) Selected features: N/A |

While performance metrics of these studies are not directly comparable due to variations in datasets and methodologies, it is noteworthy that all these approaches utilized a whole-tumor analysis strategy. In contrast, our study introduces a novel approach using super-voxel segmentation with variance-based feature aggregation. This captures localized heterogeneity within tumors and achieves an improved test AUC of 0.92.

Super-voxel segmentation divides the tumor into smaller, more homogeneous regions, which allows for a more granular analysis of each subregion. Therefore, our results showed that the subregion approach which takes into account heterogeneity, improves the HPV status prediction accuracy significantly with respect to conventional algorithms which assume that the tumor is homogenous.

Our results highlights the importance of considering tumor heterogeneity for improved predictions. Supporting this, Xie et al. [18] in esophageal cancer and Pan et al. [17] in head and neck cancer explored subregional analysis for predicting patient outcomes. Both studies showed that models based on subregion analysis performed better than those using whole-tumor approaches. Similarly, Cherezov et al. [16] showed the predictive power of subregion computation, achieving an AUC of 0.85 for lung cancer malignancy prediction. These findings further support the hypothesis that heterogeneity features provide added value over whole-tumor analysis.

### 4.1. Limitations and Future Directions

Our study has some limitations. First, the dataset size was relatively small, which may limit the generalizability. Second, our data had a class imbalance, with a higher proportion of HPV-positive cases compared to HPV-negative cases. This imbalance may have affected the performance of the predictive models, even though we have used an appropriate resampling method to counter the effect of imbalanced data. In future work, applying our model to larger datasets with more balanced class distributions will be important, as there is no universally optimal rule for addressing class imbalance [37]. Additionally, the choice of feature selection methods could have influenced our results. Future studies could investigate the impact of different feature selection strategies on the model’s performance to further optimize and validate our approach.

In future studies, we aim to explore the biological relevance of tumor subregions. Furthermore, we plan to implement a population level habitat analysis framework to further refine our model. We may uncover biological mechanisms underlying habitat imaging subtypes by exploring tumor habitats through this approach.

## 5. CONCLUSION

In conclusion, the subregion approach which takes into account heterogeneity, improves the HPV status prediction accuracy significantly with respect to conventional algorithms which assume that the tumor is homogenous. This contribution may facilitate further the use of radiomics for HPV status prediction in clinical practice. This study also confirms from the radiomics point of view that HPV positive tumors display higher levels of homogeneity. Future work will expand the analysis to a multicentered dataset to strengthen these findings and improve the robustness of our conclusion.

## Data Availability

All data produced in the present study are available upon reasonable request to the authors.

## Notes

### Competing Interest Statement

The authors have declared no competing interest.

### Funding Statement

Oya Altinok gratefully acknowledges the support provided by the Fulbright Visiting Scholar Program for this research.

### Author Declarations

The study used publicly available data obtained from The Cancer Imaging Archive (TCIA). Access to the dataset required agreeing to the TCIA Restricted License Agreement. RESTRICTED LICENSE AGREEMENT of THE CANCER IMAGING ARCHIVE gave approval for this work. The data can be accessed at: https://wiki.cancerimagingarchive.net/display/DOI/Radiomics+outcome+prediction+in+Oropharyngeal+cancer#33948240036220c66a5a436f90e4a0b54367bfae

